# Cardiac Stasis Imaging, Stroke and Silent Brain Infarcts in Patients with Non-Ischemic Dilated Cardiomyopathy

**DOI:** 10.1101/2024.03.22.24304765

**Authors:** Elena Rodríguez-González, Pablo Martínez-Legazpi, Ana González-Mansilla, M. Ángeles Espinosa, Teresa Mombiela, Juan A. Guzmán-De-Villoria, Maria Guadalupe Borja, Fernando Díaz-Otero, Rubén Gómez de Antonio, Pilar Fernández-García, Ana I Fernández-Ávila, Cristina Pascual-Izquierdo, Juan C del Álamo, Javier Bermejo

## Abstract

**Background:** Cardioembolic stroke is one of the most devastating complications of non-ischemic dilated cardiomyopathy (NIDCM). However, in clinical trials of primary prevention, the benefits of anticoagulation were hampered by the risk of bleeding. If indices of cardiac blood stasis account for the risk of stroke, they may be useful to individualize primary prevention treatments.

**Methods:** We performed a cross-sectional study in patients with NIDCM and no history of atrial fibrillation (AF) from two sources: 1) a prospective enrollment of unselected patients with left ventricular (LV) ejection fraction <45% and 2) a retrospective identification of patients with a history of previous cardioembolic neurological event. The primary endpoint integrated a history of ischemic stroke, transient ischemic attack (TIA), or the presence intraventricular thrombus, or a silent brain infarction (SBI) by imaging. From echocardiography, we calculated blood flow inside the LV and its residence time (*R_T_*). The study was registered in ClinicalTrials.gov (NCT03415789).

**Results:** Of the 89 recruited patients, 18 showed a positive primary endpoint: 9 patients had a history stroke or TIA and another 9 were diagnosed with SBIs in the brain imaging. *R_T_* performed good to identify the primary endpoint (AUC (95% CI)= 0.75 (0.61-0.89), p= 0.001). A *R_T_* > 2.21 cycles showed a sensitivity of 0.88 (0.77-1.00) and specificity of 0.70 (0.10-0.81). When accounting only for identifying a history of stroke or TIA, AUC for *R_T_* was 0.92 (0.85-1.00) with and odds ratio= 7.2 (2.3 – 22.3) per cycle, p< 0.001.

**Conclusions:** In patients with NIDCM in sinus rhythm, stasis imaging derived from echocardiographyis is closely related to the burden of stroke. Stasis imaging may be useful to address stroke risk in patients with systolic dysfunction.

## Introduction

Cardioembolic stroke is associated with a high in-hospital mortality.^1^ Around 20% of these strokes occur in patients with sinus rhythm and structural heart disease, and left ventricular (LV) systolic dysfunction, due to either ischemic or nonischemic dilated cardiomyopathy (NIDCM), is the most frequent finding in these patients.^2^ In fact, patients with NIDCM show an annual incidence of stroke 3 to 8 times higher than the age-matched population.^3^ Several studies have also demonstrated a high prevalence of silent brain infarction (SBI) in the setting of dilated cardiomyopathy, even in absence of atrial fibrillation (AF).^4^ SBIs are a documented source of disability and mortality, and both their presence and number are highly sensitive predictors of clinical stroke.^5,6^

Randomized controled clinical trials have failed to demonstrate a significant benefit for oral anticoagulation in non-AF patients with heart failure (HF) and low ejection fraction, as the reduction of thrombotic events was counterbalanced by the incidence of bleeding complications.^7^ Trials aimed to secondary prevention after embolic strokes of unknown source have also been neutral.^8^ Therefore, there is an unmet medical need for developing robust markers to address the risk of cardioembolism in non-AF HF patients.

In NIDCM, global and regional LV chamber abnormalities induce an abnormal blood flow transit inside the chamber that leads to regions of increased blood stasis.^9^ Combined with the prothrombotic state caused by HF neurohormonal activation, stasis is a risk factor for cardioembolic events, even in absence of AF.^10^ In recent years, computational image methods to quantify blood stasis inside the heart using conventional modalities have been developed.^11,12^ In this regard, an ultrasound-based biomarker of stasis, based on the number of cycles spent by blood particles inside the LV (i.e., the *residence time R_T_*), has been proposed. Preliminary studies have shown the potential of *R_T_* for addressing the risk of cardioembolism in patients with ventricular dysfunction after STEMI.^13–15^ This index has also been suggested to be a biomarker of the significance of coronary artery stenosis, atherosclerosis^16^ or ventricular dysfunction.^17^ In this study, we aimed to assess the performance of stasis imaging to account for the burden of cardioembolic stroke in NIDCM.

## Methods

We designed the ISBIDCM clinical trial, “Imaging Silent Brain Infarct in Dilated Cardiomyopathy” (NCT 03415789, first posted January 2018), to decipher the relationship of intraventricular stasis with the prevalence of SBIs, intraventricular mural thrombosis and/or stroke in patients with NIDCM. Inclusion criteria were the diagnosis of NIDCM with a LV ejection fraction ≤ 45% (unrelated to ischemic cardiomyopathy), age ≥ 18 years old and no history of AF. Exclusion criteria were any contraindication for MR examination, any primary valve disease ≥ 3+ severity, a diagnosis of carotid artery disease with a > 50% stenosis, oral anticoagulation before enrollment, history of prothrombotic disease, and reluctance to sign the written informed consent.

The sample size for the ISBIDCM study was established based on an estimated prevalence of SBIs of 30-35%.^4^ However, after achieving the recruitment target, the number of patients with the primary endpoint was 11% (see below). As this markedly lowered the power of the study, the Steering Committee approved enriching the positive endpoint population with an additional group of NIDCM patients who had a medical history of ischemic neurological events and met the other inclusion criteria for the ISBIDCM study. Therefore, we added additional cases to the study by retrospectively screening patients from two sources: 1) all admissions to the emergency department from Nov-2019 to Feb-2022 with ICD-10 codes of ischemic stroke or transient ischemic attack (TIA), who received a diagnosis of NIDCM during cardiac workup, and 2) all patients with NIDCM followed in the cardiomyopathy clinic with a history of stroke or transient ischemic attack. Any history of AF (either clinically or sub-clinically detected in Holter tests or by any cardiac monitoring device) was an exclusion criterion for both sources. Thus, the present report comprises both the original ISBIDCM cohort and this additional group of patients with a previous history of stroke or TIA (**Figure 1**). The Ethics Committee of the Hospital Gregorio Marañón approved the study and all patients provided written informed consent. The study was academically funded by a grant of Spanish Cardiology Society, and the Fundación para la Investigación Biomédica Hospital Gregorio Marañón was the unique sponsor. All patients underwent an exhaustive echocardiography. In patients with no history of ischemic neurological event a cardiac and brain magnetic resonance examinations and blood test were also performed.

**FIGURE 1:**
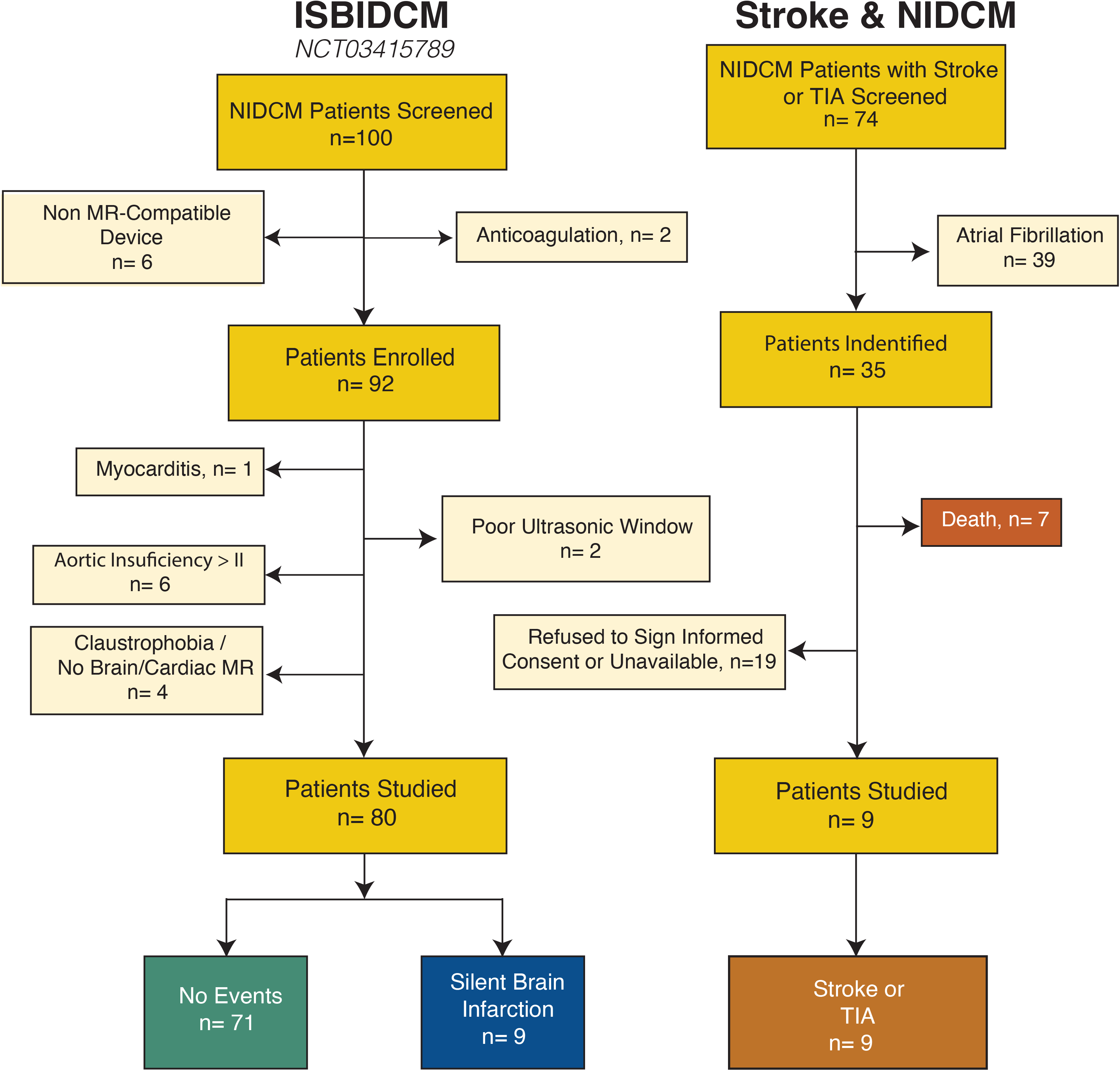
Flow Diagram of Patient Populations and Primary Endpoints. TIA: transient ischemic attack

### Study Endpoints

The primary endpoint for the study was either: 1) the identification of an any-date SBI lesion in the brain MR examination, 2) the presence of an intraventricular thrombus in any imaging test, or 3) a clinical history of stroke or TIA (**Figure 1**).

### Ancillary Variables and Blood Biomarkers

Clinical data included past medical history and ongoing medications, as well as hematological and biochemical assessments obtained from medical records within the 2 months before or after inclusion. In all patients of the original cohort a comprehensive assessment of coagulation, fibrinolysis biomarkers, von Willebrand Factor and ADAMTS13 activity was performed, and they underwent Beck Depression Inventory and the Mini-Mental State Examinations to screen for depression and dementia, respectively. Transcranial Doppler and carotid duplex ultrasonographic examinations, and 24-hour Holter monitoring test were performed in patients with the primary endpoint to rule-out alternative causes of stroke, TIA, or SBI. CHA_2_DS_2_-VASc score was calculated.

### Cardiac Imaging

Echocardiographic examinations were performed at the enrollment using a Vivid 7 scanner and a phase-array 2-4 MHz transducer (GE Healthcare). We obtained 3-dimensional apical sequences to measure LV volumes and ejection fraction (EF). Longitudinal myocardial strain was measured from apical long-axis sequences using a 6-segment model (EchoPac version 204, GE Healthcare). The E-wave propagation index, a reported biomarker of mural trhonbosis, was computed as the E-wave velocity time integral over the LV long-axis length. The CMR imaging protocol included a cine steady-state free precession imaging of LV function (SENSE X 2, repetition time: 2.4 ms, echo time: 1.2 ms, average in-plane spatial resolution: 1.6 x 2 mm, 30 phases per cycle, 8-mm slice thickness without gap) and late enhancement imaging (3D inversion-recovery turbo gradient echo sequence, pre-pulsed delay optimized for maximal myocardial signal suppression; 5-mm actual slice thickness, inversion time: 200-300 and 600 ms). Images were obtained in short axis (10 to 14 contiguous slices) and 4-, 2-, and 3-chamber views. Late enhancement sequences were obtained 3 to 10 min after injection of 0.1 mmol/kg of gadobenate dimeglumine (ProHance, Bracco Imaging) to assess for mural thrombosis and quantify myocardial focal fibrosis.

### Brain Imaging

Brain MR was performed along CMR imaging and included sagittal T1-weighted images, axial diffusion weighted images, coronal T2-weigthed turbo spin echo (TSE) and axial FLAIR-T2-weigthed images. Acute or subacute diffusion weighted image strokes were adjudicated whenever focal lesions greater than 3 mm hyperintense on diffusion weighted images with low apparent coefficient diffusion or pseudonormalization values were respectively identified. Chronic ischemic injury was diagnosed when the lesion was isointense to CSF on T2 and FLAIR weighted images appeared surrounded by an hyperintense lineal rim of gliosis and showed high apparent coefficient diffusion values. All studies were interpreted by expert neuroradiologists who also differentiated these types of lesions from dilated perivascular spaces based on their distribution and morphology

### Intraventricular Stasis Imaging

From Color-Doppler, we obtained 2-dimensional, time-resolved (2D+t) blood flow velocity vector fields in the apical long-axis view of the LV, as described elsewhere.^18^ The 2D+t blood flow fields were integrated using a forced advection equation for mapping and quantifying the residence time, *R_T_*, as previously reported.^11,13^ We computed the evolution of *R_T_* for 8 consecutive cardiac cycles, the estimated period taken for a full washout in a normal LV.^19^ We designed a color-coded scale to generate video maps of RT of blood in the LV. In each frame, the color scale represents the number of cardiac cycles a blood particle has spent inside the chamber; dark blue represents “fresh” blood recently entering the LV, whereas dark red represents stasis regions in which with blood is retained for at least 4 cardiac cycles (**see Videos** for details). At the end of the 8^th^ beat, we collected the average *R_T_* inside the LV as a single scalar. The reproducibility of *R_T_* has been reported elsewhere.^19^

### Statistical Analysis

Data are described as median [interquartile range] except otherwise indicated. We used Wilcoxon rank-sum, Chi-squared and Fisher exact tests to compare quantitative variables and proportions, respectively. We used univariate and bivariate logistic regression models to address the identification of the primary endpoint, adjusting for clinical factors (see below). Odds-ratios and their 95% confidence intervals (CIs) are reported for these models. The interplay between clinical variables, *R_T_*, and the primary endpoint was further investigated using mediation analysis. To test for diagnostic performance, we used ROC analyses to compute the area under the curve (AUC), its 95% CI, and its statistical significance. Cutoffs were selected by the Youden’s method, weighted to penalize for false negatives. Medians (and 95% CIs) of performance metrics of these cutoffs (sensitivity, specificity) were calculated by bootstrapping with 2,000 replicates. All statistical analyses were performed in R (v. 4.1.3) and p-values < 0.05 were considered significant.

## Results

We included 89 patients (80 patients from the original ISBIDCM cohort and 9 patients with diagnosis of stroke or ischemic transient attack from the additional cohort), with a median age of 59 [49-69] years old, and 37 (42%) were women. The primary endpoint was recorded in 18 patients (20%): 9 patients with a history of stroke or TIA, and 9 with identified SBIs (2 patients with 3 lesions, 1 patient with 2, and 6 patients with 1); no patient was identified with an intraventricular thrombus. A total of 55% of the SBI were found in the cerebellum, 3 of them were located cortically, and one of the events had cortico-subcortical location. Carotid duplex and transcranial Doppler ruled out alternative etiologies of the neurological events in all patients with a primary endpoint.

There were no differences in age, sex and other clinical variables as NYHA class or current treatment among patients with and without the primary endpoint. However, patients with a positive primary endpoint showed more frequently diabetes mellitus and dyslipidemia, (53 vs. 27% and 63 vs. 35%, respectively: all p< 0.05) and had higher CHA2DS2-VASc scores (3 [3-5] vs 2 [2-3], p=0.004) than those without an endpoint. Also, patients with stroke or SBI showed higher levels of creatinine (1.20 [0.88-1.42] vs 0.87 [0.77-1.08] mg/dL, p= 0.01). Other hematological, coagulation and biochemical variables were similar in both groups (**Table 1**). Patients with a primary endpoint showed lower Mini-Mental scores than patients without one, whereas there were no differences in the Beck Depression Test.

**Table 1.**
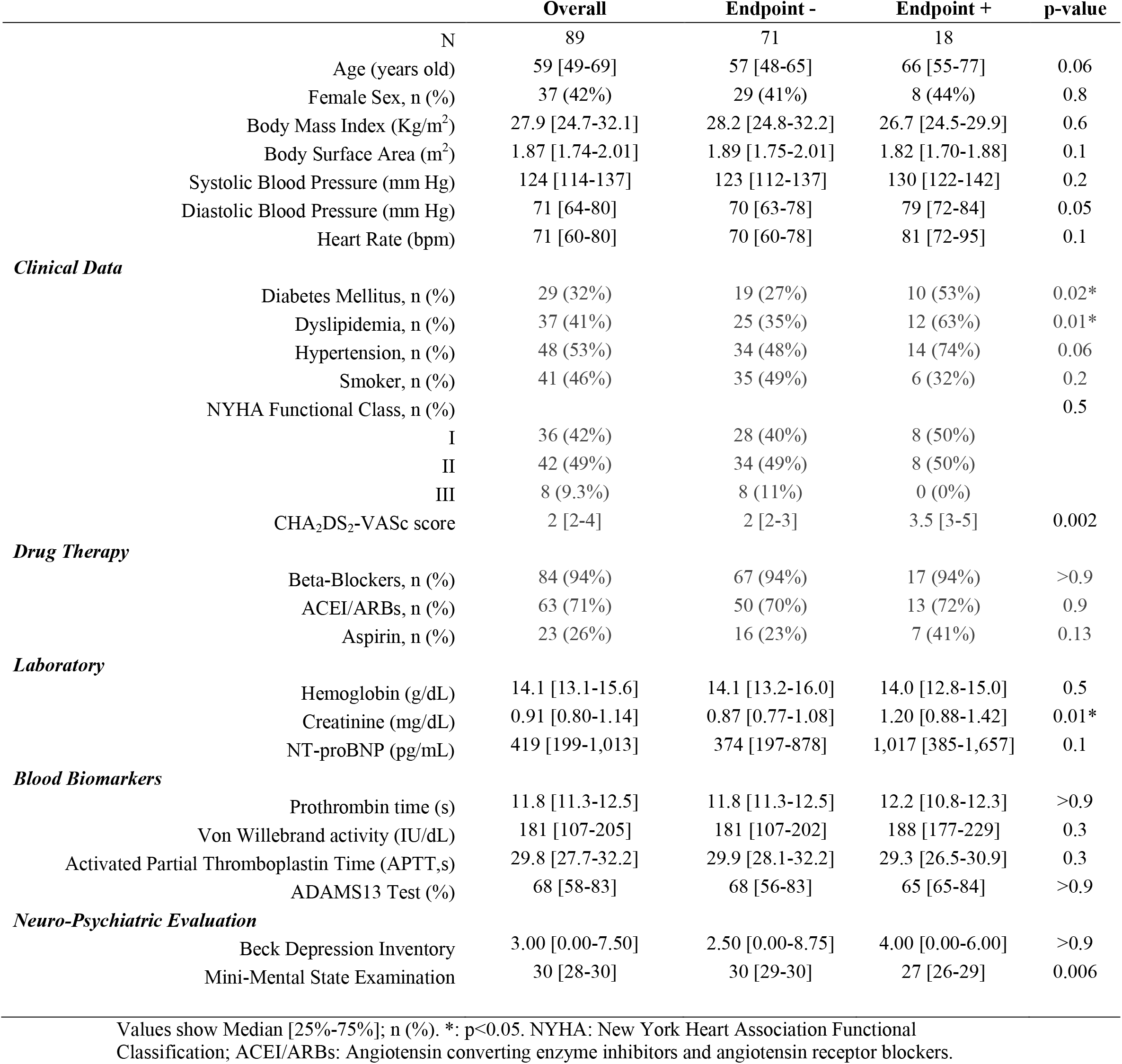
Clinical Data.

|  | Overall | Endpoint - | Endpoint + | p-value |
| --- | --- | --- | --- | --- |
| N | 89 | 71 | 18 |  |
| Age (years old) | 59 [49-69] | 57 [48-65] | 66 [55-77] | 0.06 |
| Female Sex, n (%) | 37 (42%) | 29 (41%) | 8 (44%) | 0.8 |
| Body Mass Index (Kg/m<sup>2</sup>) | 27.9 [24.7-32.1] | 28.2 [24.8-32.2] | 26.7 [24.5-29.9] | 0.6 |
| Body Surface Area (m<sup>2</sup>) | 1.87 [1.74-2.01] | 1.89 [1.75-2.01] | 1.82 [1.70-1.88] | 0.1 |
| Systolic Blood Pressure (mm Hg) | 124 [114-137] | 123 [112-137] | 130 [122-142] | 0.2 |
| Diastolic Blood Pressure (mm Hg) | 71 [64-80] | 70 [63-78] | 79 [72-84] | 0.05 |
| Heart Rate (bpm) | 71 [60-80] | 70 [60-78] | 81 [72-95] | 0.1 |
| **Clinical Data** |  |  |  |  |
| Diabetes Mellitus, n (%) | 29 (32%) | 19 (27%) | 10 (53%) | 0.02* |
| Dyslipidemia, n (%) | 37 (41%) | 25 (35%) | 12 (63%) | 0.01* |
| Hypertension, n (%) | 48 (53%) | 34 (48%) | 14 (74%) | 0.06 |
| Smoker, n (%) | 41 (46%) | 35 (49%) | 6 (32%) | 0.2 |
| NYHA Functional Class, n (%) |  |  |  | 0.5 |
| I | 36 (42%) | 28 (40%) | 8 (50%) |  |
| II | 42 (49%) | 34 (49%) | 8 (50%) |  |
| III | 8 (9.3%) | 8 (11%) | 0 (0%) |  |
| CHA<sub>2</sub>DS<sub>2</sub>-VASc score | 2 [2-4] | 2 [2-3] | 3.5 [3-5] | 0.002 |
| **Drug Therapy** |  |  |  |  |
| Beta-Blockers, n (%) | 84 (94%) | 67 (94%) | 17 (94%) | >0.9 |
| ACEI/ARBs, n (%) | 63 (71%) | 50 (70%) | 13 (72%) | 0.9 |
| Aspirin, n (%) | 23 (26%) | 16 (23%) | 7 (41%) | 0.13 |
| **Laboratory** |  |  |  |  |
| Hemoglobin (g/dL) | 14.1 [13.1-15.6] | 14.1 [13.2-16.0] | 14.0 [12.8-15.0] | 0.5 |
| Creatinine (mg/dL) | 0.91 [0.80-1.14] | 0.87 [0.77-1.08] | 1.20 [0.88-1.42] | 0.01* |
| NT-proBNP (pg/mL) | 419 [199-1,013] | 374 [197-878] | 1,017 [385-1,657] | 0.1 |
| **Blood Biomarkers** |  |  |  |  |
| Prothrombin time (s) | 11.8 [11.3-12.5] | 11.8 [11.3-12.5] | 12.2 [10.8-12.3] | >0.9 |
| Von Willebrand activity (IU/dL) | 181 [107-205] | 181 [107-202] | 188 [177-229] | 0.3 |
| Activated Partial Thromboplastin Time (APTT,s) | 29.8 [27.7-32.2] | 29.9 [28.1-32.2] | 29.3 [26.5-30.9] | 0.3 |
| ADAMS13 Test (%) | 68 [58-83] | 68 [56-83] | 65 [65-84] | >0.9 |
| **Neuro-Psychiatric Evaluation** |  |  |  |  |
| Beck Depression Inventory | 3.00 [0.00-7.50] | 2.50 [0.00-8.75] | 4.00 [0.00-6.00] | >0.9 |
| Mini-Mental State Examination | 30 [28-30] | 30 [29-30] | 27 [26-29] | 0.006 |

Values show Median [25%-75%]; n (%). \*: p<0.05. NYHA: New York Heart Association Functional Classification; ACEI/ARBs: Angiotensin converting enzyme inhibitors and angiotensin receptor blockers.

Traditional cardiac imaging indices showed no significant differences among patients with and without the primary endpoint. In fact both groups showed nearly identical values of all systolic and diastolic variables (**Table 2**). Left atrial diameter was similar in both groups (4.1 [3.6-4.5] vs. 4.0 [3.6-4.6] cm, p=0.9), and no differences were found in focal fibrosis as detected by LGE in MR. Also, the global and apical LV strains measured by echocardiography were similar in patients with and without the endpoint.

**Table 2:**
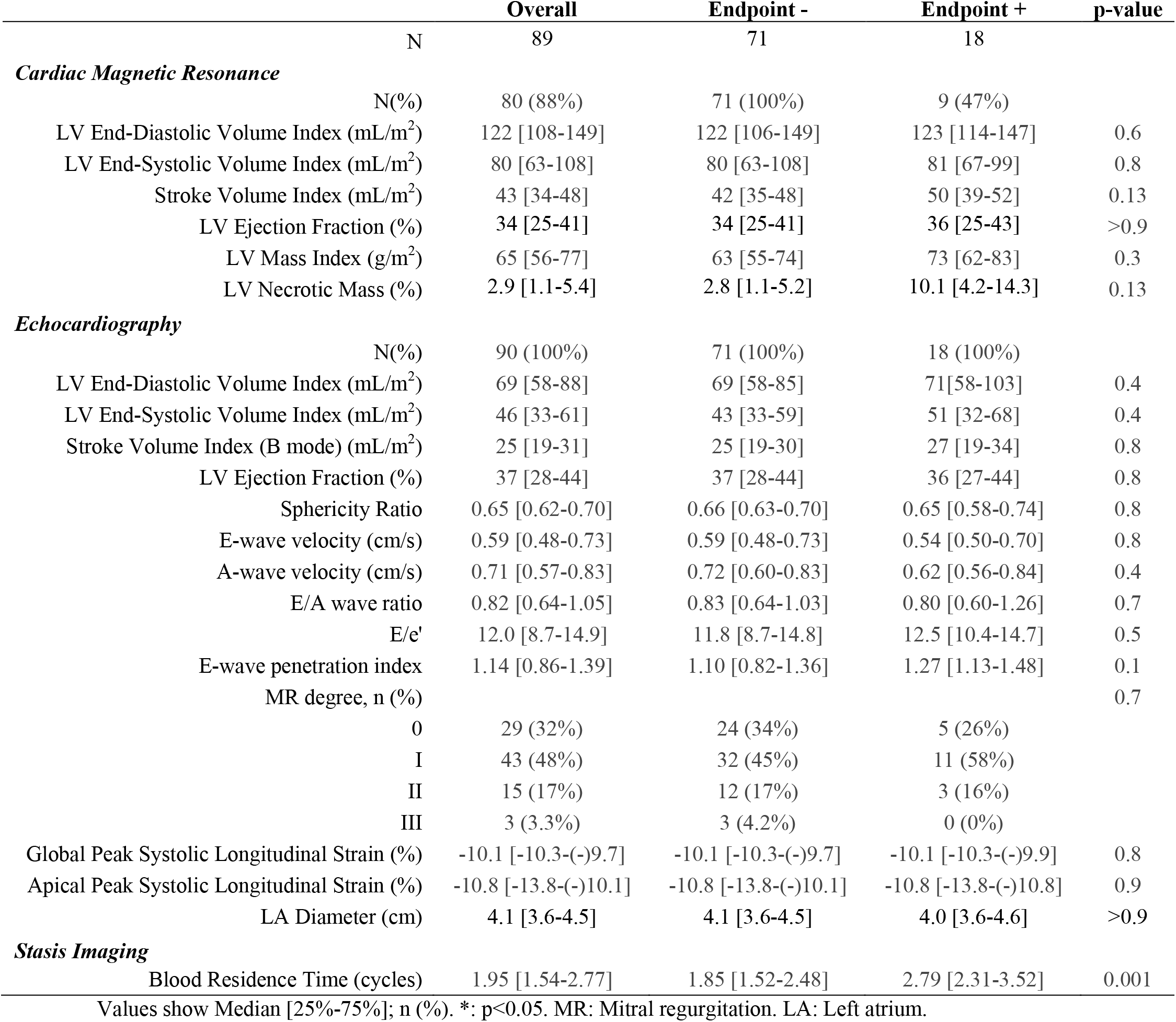
Cardiac Imaging.

### Stasis Imaging & Primary Endpoint

*R_T_* performed favourably to identify the primary endpoint (AUC = 0.75, 95% CI= 0.61-0.89, OR= 2.83 per cardiac cycle, 95% CI= 1.50 – 5.34, p= 0.001). Furthermore, *R_T_* remained significantly associated to the primary endpoint in bivariate models adjusted for covariables such as diabetes, dyslipidemia, hypertension, and creatinine levels (OR in the range of 2.51-2.74, all p< 0.01). The increased risk related to these clinical risk factors was not mediated by a prolongation of *R_T_*(p> 0.8 for all mediation effects). The threshold *R_T_*> 2.21 cycles showed a sensitivity of 0.88 (0.77-1.00), and a specificity of 0.71 (0.10-0.81). When accounting only for the relationship with stroke or TIA, the AUC for *R_T_*was 0.92 (0.85-1.00) with an OR= 7.2, 95% CI= 2.30 – 22.3, p< 0.001 (**Figure 2**).

**FIGURE 2:**
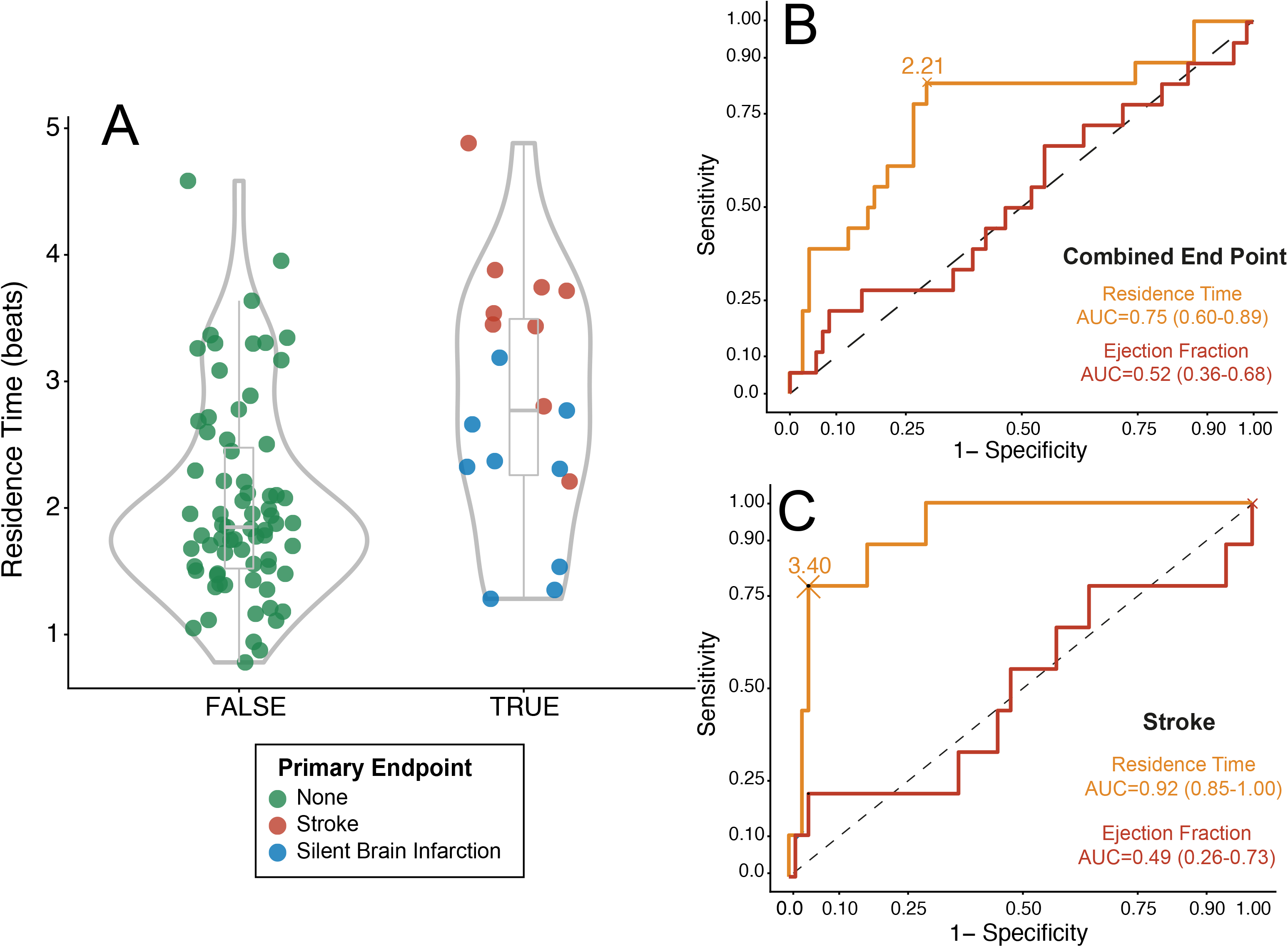
Relationship between Residence Time and the Primary Endpoint. **Panel A.** Violin plot of the distributions of residence time (*R_T_*). The primary endpoint is colored upon its etiology. **Panel B:** Receiver operating characteristic curve for the performance of *R_T_* for addressing the primary endpoints. **Panel C:** Receiver operating characteristic curve for the performance of *R_T_*for assessing stroke.

The overall relationship the primary endpoint with *R_T_* was higher than that other conventional imaging parameters, such as LV EF [AUC= 0.52 (0.36-0.68)], global and apical strain (AUCs of 0.44 and 0.55, respectively, p> 0.05 for both), and E-wave penetration index (AUC of 0.62, p= 0.07).^20^ **Figure 3** shows illustrative examples of stasis imaging and cardiac and brain magnetic resonance for a patient with stroke, a patient with SBIs and a patient free of events.

**FIGURE 3:**
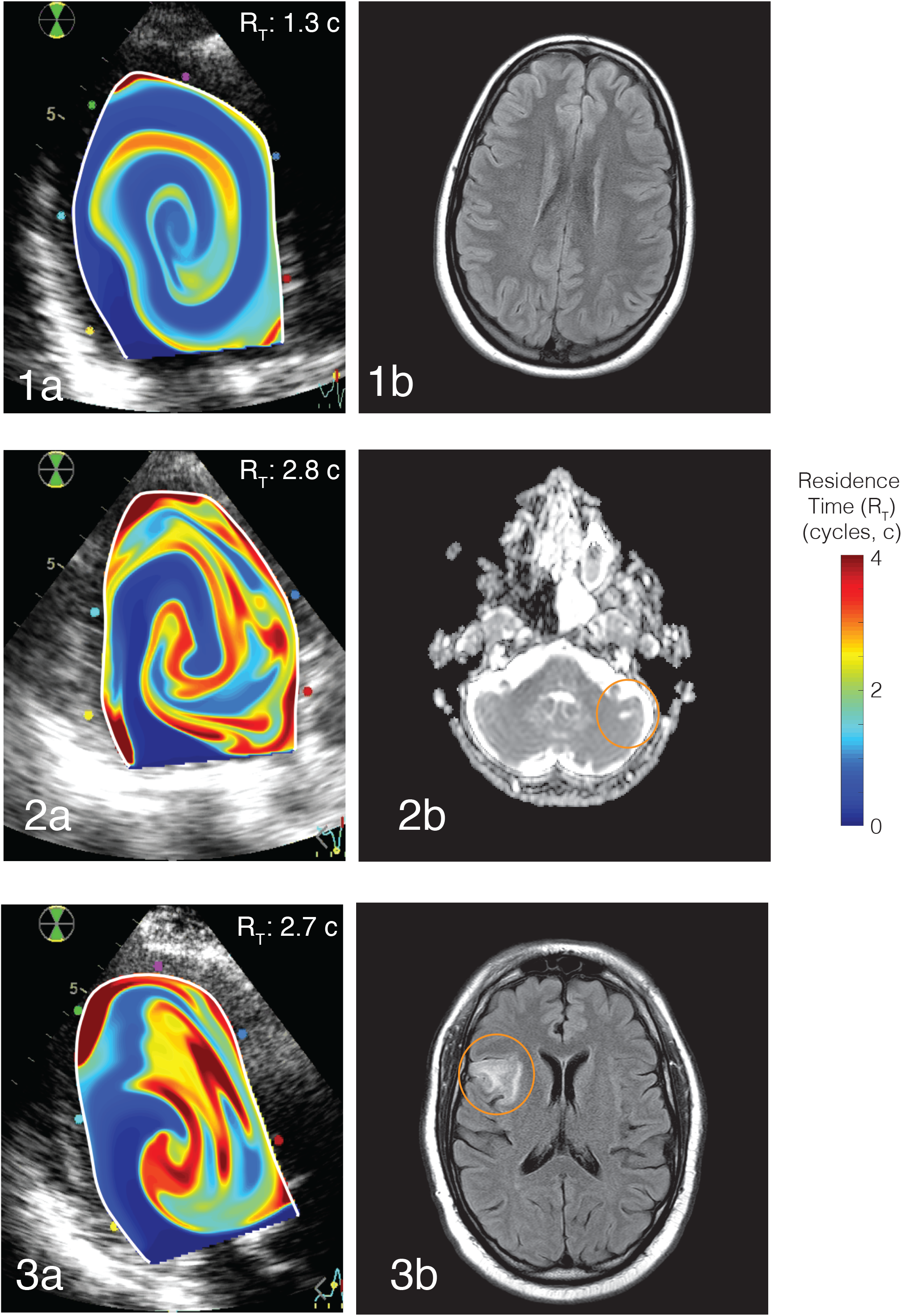
Examples of Residence Time Mapping and Brain MRI. **Panel 1 (a-b)**: *R_T_* distribution in the LV and brain MR of a NIDCM patient without event. **Panels 2 & 3 (a-b)**: *R_T_* distribution in the LV and brain MR of two NIDCM patients with a SBI (**Panel 2)** and a stroke (**Panel 3)**. *R_T_* maps are overlaid on the B-mode echocardiogram.

## Discussion

As far as we know, this is the first study aimed to validate an imaging-based biomarker of intracardiac stasis against the burden of brain infarcts, TIAs and stroke in a moderately large cohort of patients with non-ischemic dilated cardiomyopathy, in the absence of AF. Classic imaging markers of intraventricular thrombosis such as EF were not related to the burden of cardioembolic stroke in our cohort. Moreover, advanced imaging indices, such as longitudinal strain or E-wave penetration index, which have been proven useful to predict thrombosis in the LA in patients with AF,^21^ or after STEMI in the LV,^15,20,22^ also showed useless relationship with cardioembolic burden in our study. In contrast, the residence time of blood from echocardiographic vector flow mapping was closely related to ischemic brain events, whether clinical or silent. This biomarker may be useful to personalize treatment on those scenarios were indiscriminate anticoagulation is not supported by evidence.^23^

### Silent Brain Infarct in NIDCM

SBIs are defined as those brain lesions that meet characteristics of an ischemic event in the absence of neurological symptoms or a previous history of infarction or transient ischemic attack.^24^ SBIs are a well-known risk marker for both clinical stroke^25^ and global mortality in population studies,^26^ and are the cause of significant disability linked to cognitive impairment, dementia, and depression.^27^ Left ventricular thrombus formation, cardiomyopathy, atrial fibrillation and patent foramen oval have all been associated with SBIs^28^ and they have been considered precursors of ischemic stroke.

In patients with NIDCM, prevalence of SBIs has been disclosed to be as high as 35%,^4^ and intraventricular thrombosis occurrence has been reported to be around 13%, frequently accompanied by high prevalence of thrombi in the atrium and left atrial appendage. In our prospective study group, the prevalence of SBI was 10%, and we did not observe LV mural thrombosis in any patient. This prevalence is lower than previously reported, probably because previous studies included patients with ischemic cardiomyopathy, AF,^29^ and lower ejection fraction.^29–31^ The fact that we did not identify any case of mural thrombosis by LGE-CMR, does not exclude a cardioembolism as the etiology of the ischemic events, as the stagnant regions in the cardiac chambers may activate the coagulation cascade without macroscopic thrombosis ever being detected, either because a small mural thrombus is missed or because all the thrombotic material embolizes before the patient is scanned.

### Intraventricular blood stasis as a source of cardioembolism in NIDCM

As one of the three pillars of Virchow’s triad, blood stasis is a recognized key factor for cardiac embolism.^32^ However, until recently, the assessment of blood stasis inside the heart was qualitatively based only on the visual interpretation of spontaneous contrast from B-mode ultrasound. Although such contrast is related to the risk of thrombosis both in the left atrium^33^ and the LV,^4^ it is highly dependent on equipment, operator, and factors such as the relative concentrations of red blood cells and fibrinogen.^34^

Advances in medical imaging now make it possible to measure time-resolved two-or even three-dimensional flow velocity fields inside the cardiac chambers.^35^ The analysis of these datasets using the laws of fluid mechanics allows for quantifying flow-related indices and to derive metrics accounting for the transport and stagnation of blood inside the cardiac chambers.^36,37^ One of these indices, the residence time, *R_T_*, is particularly well-suited to quantify blood stasis, as it can measure the time that fluid particles spend inside a cardiac chamber.^11,38^ The performance of *R_T_* to predict cardioembolism has been recently demonstrated in the setting of STEMI. In a porcine model, high *R_T_* in the LV has been shown to correlate with the occurrence of high-intensity signals suggestive of cerebral microembolism in carotid Doppler ultrasound scans.^14^ In patients with STEMI, *R_T_* has also shown good performance to predict intraventricular thrombus, SBI and stroke.^15^ The present study significantly extends these previous findings by confirming that *R_T_* may also be a good biomarker of SBI and stroke beyond the specifics found in the infarcted LV.

### Flow patterns and stasis in NIDCM patients

The diastolic LV swirling flow pattern routes blood entering the chamber to displace the blood volume lingering from previous beats towards the outflow tract,^39^ helping to minimize the residence time of blood. The changes in chamber size and morphology associated with NIDCM strengthen this pattern, creating larger, more persistent swirling structures as compared to normal hearts^9^ (see an example in **Figure 4, Videos 1 & 2**). This mechanism could at least partially compensate the increase in *R_T_* concomitant with reduced EF, and differences in flow patterns among patients would explain why EF is not a good predictor of *R_T_* and adverse events in our cohort.^40,41^

**FIGURE 4:**
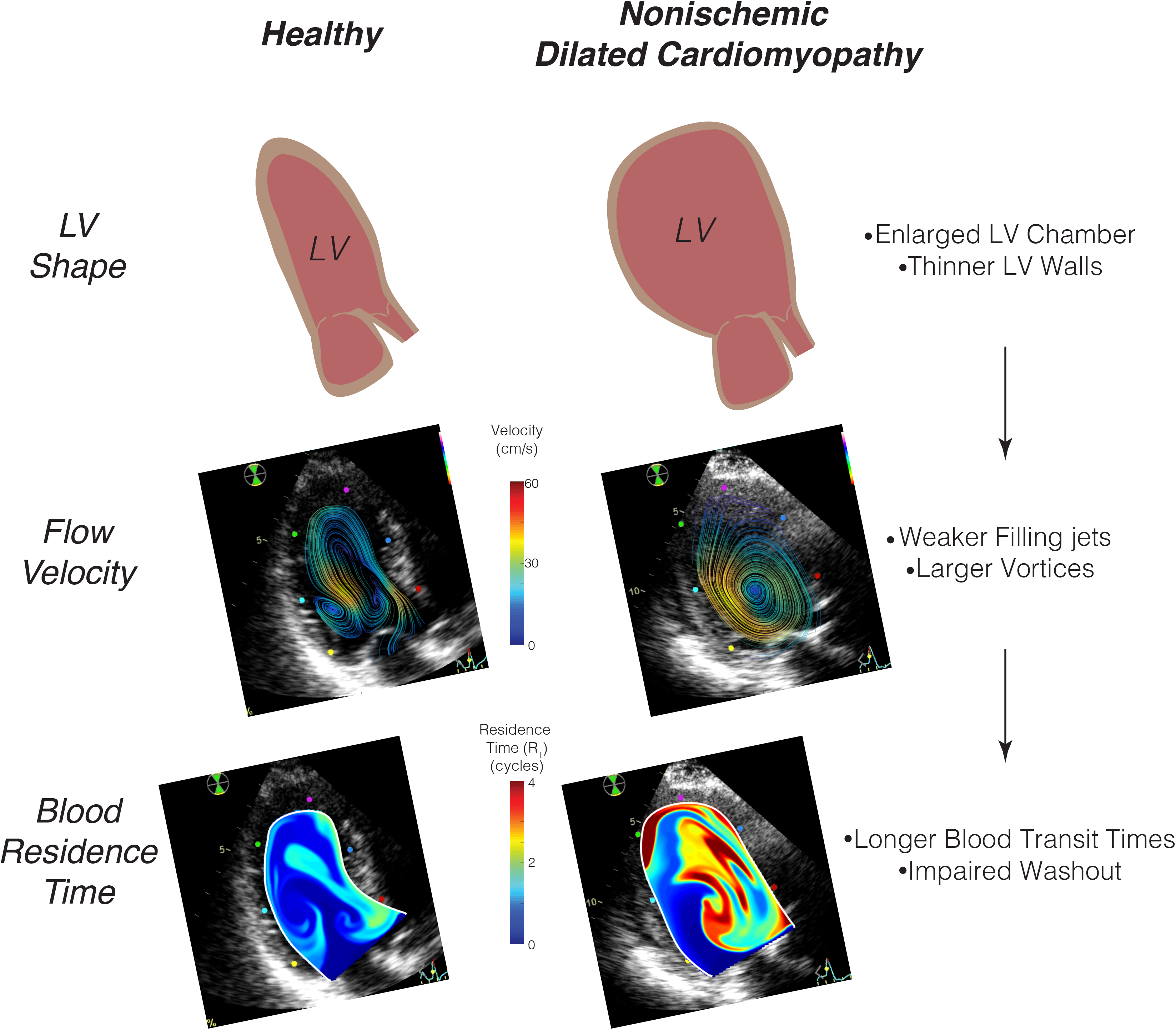
Intraventricular flow and Residence time in the normal heart (left) and in the patients with NIDCM (right).

### Limitations

This was a monocentric study, and the number of events was small. Therefore, further validation in larger multicentric clinical trials need to be conducted. Alternative etiologies may result in SBIs beyond cardioembolism; to reduce the possibility of secondary sources, we performed comprehensive etiological research in all patients with SBIs and excluded all patients with any history of known or suspected AF. However, small vessel disease or subclinical AF were impossible to address by study design and could be responsible for some of the identified lesions. The hemodynamic status of each individual patient may have changed during the occurrence of the event and the scanning time and therefore, the association between residence time and stroke shall be further addressed in prospective trials. Stasis imaging was based on conventional 2D transthoracic echocardiography, assuming a planar-flow distribution inside the LV. Although this is an inherent methodological simplification, several studies have validated this hypothesis.^42^

### Conclusions

In patients with NIDCM in sinus rhythm, cardioembolic risk may be evaluated using bedside echocardiography combined with stasis imaging. This imaging modality may be useful for addressing stroke risk in patients with NIDCM and no history of AF.

## Data Availability

Data will be available upon reasonable request to the authors.

## Acknowledgments

We are in debt with all the personnel of the Department of Cardiology of the Hospital General Universitario Gregorio Marañón.

## Video Legends

**VIDEO 1**: Residence Time evolution in a patient with stroke. The RT map is colored and overlaid over a B-mode of the 3-chamber view of the LV. Flow colors account for the time each blood volume spends inside the LV, from dark blue (blood entering in the LV) to dark red (blood spending more than 4 cycles inside the chamber). As time evolves, due to deficient blood mixing and stagnation the hue of blood shifts towards red colors.

**VIDEO 2**: Residence Time evolution in a patient free of events. The RT map is overlaid over a B-mode of the 3-chamber view of the LV. Flow colors are identical to represented in Supplemental video 1. Notice that contrary to what is shown in that video, adequate blood mixing and washout are depicted by low values of residence time.

## References

1. Collaborators GBDS. Global, regional, and national burden of stroke and its risk factors, 1990-2019: a systematic analysis for the Global Burden of Disease Study 2019. Lancet Neurol. 2021;20:795–820.

2. Pujadas Capmany R, Arboix A, Casanas-Munoz R, et al. Specific cardiac disorders in 402 consecutive patients with ischaemic cardioembolic stroke. Int J Cardiol. 2004;95:129–134.

3. Lip GY, Rasmussen LH, Skjoth F, et al. Stroke and mortality in patients with incident heart failure: the Diet, Cancer and Health (DCH) cohort study. BMJ Open. 2012;2.

4. Kozdag G, Ciftci E, Vural A, et al. Silent cerebral infarction in patients with dilated cardiomyopathy: echocardiographic correlates. Int J Cardiol. 2006;107:376–381.

5. Bernick C, Kuller L, Dulberg C, et al. Silent MRI infarcts and the risk of future stroke: the cardiovascular health study. Neurology. 2001;57:1222–1229.

6. Fanning JP, Wesley AJ, Wong AA, et al. Emerging spectra of silent brain infarction. Stroke. 2014;45:3461–3471.

7. Beggs SAS, Rorth R, Gardner RS, et al. Anticoagulation therapy in heart failure and sinus rhythm: a systematic review and meta-analysis. Heart. 2019;105:1325–1334.

8. Ntaios G, Vemmos K, Lip GY. Oral anticoagulation versus antiplatelet or placebo for stroke prevention in patients with heart failure and sinus rhythm: Systematic review and meta-analysis of randomized controlled trials. Int J Stroke. 2019;14:856–861.

9. Bermejo J, Benito Y, Alhama M, et al. Intraventricular vortex properties in non-ischemic dilated cardiomyopathy. Am J Physiol Heart Circ Physiol. 2014;306:H718–729.

10. Lip GY GC. Does heart failure confer a hypercoagulable state? Virchow’s triad revisited. Jou Amer Coll Card. 1999;33:1424–1142.

11. Rossini L, Martinez-Legazpi P, Vu V, et al. A clinical method for mapping and quantifying blood stasis in the left ventricle. J Biomech. 2016;49:2152–2161.

12. Seo JH, Abd T, George RT, et al. A coupled chemo-fluidic computational model for thrombogenesis in infarcted left ventricles. Am J Physiol Heart Circ Physiol. 2016;310:H1567–1582.

13. Martinez-Legazpi P, Rossini L, Perez Del Villar C, et al. Stasis mapping using ultrasound: A prospective study in acute myocardial infarction. JACC Cardiovasc Imaging. 2018;11:514–515.

14. Delgado-Montero A, Martinez-Legazpi P, Desco MM, et al. Blood stasis imaging predicts cerebral microembolism during acute myocardial infarction. J Am Soc Echocardiogr. 2020;33:389–398.

15. Rodríguez-González E, Martínez-Legazpi P, Mombiela T, et al. Stasis Imaging Predicts the Risk of Cardioembolic Stroke Related to Acute Myocardial Infarction. medrxiv. 2023:2023.2009.2015.23295650.

16. Hashemi J, Rai S, Ghafghazi S, et al. Blood residence time to assess significance of coronary artery stenosis. Sci Rep. 2020;10:11658.

17. Costello BT, Qadri M, Price B, et al. The ventricular residence time distribution derived from 4D flow particle tracing: a novel marker of myocardial dysfunction. Int J Cardiovasc Imaging. 2018;34:1927–1935.

18. Garcia D, Del Alamo JC, Tanne D, et al. Two-dimensional intraventricular flow mapping by digital processing conventional color-Doppler echocardiography images. IEEE Trans Med Imaging. 2010;29:1701–1713.

19. Benito Y, Martinez-Legazpi P, Rossini L, et al. Age-dependence of flow homeostasis in the left ventricle. Front Physiol. 2019;10:485.

20. Harfi TT, Seo JH, Yasir HS, et al. The E-wave propagation index (EPI): A novel echocardiographic parameter for prediction of left ventricular thrombus. Derivation from computational fluid dynamic modeling and validation on human subjects. Int J Cardiol. 2017;227:662–667.

21. Costa C, Gonzalez-Alujas T, Valente F, et al. Left atrial strain: a new predictor of thrombotic risk and successful electrical cardioversion. Echo Res Pract. 2016;3:45–52.

22. Olsen FJ, Pedersen S, Galatius S, et al. Association between regional longitudinal strain and left ventricular thrombus formation following acute myocardial infarction. Int J Cardiovasc Imaging. 2020;36:1271–1281.

23. Zannad F, Anker SD, Byra WM, et al. Rivaroxaban in Patients with Heart Failure, Sinus Rhythm, and Coronary Disease. N Engl J Med. 2018;379:1332–1342.

24. Wardlaw JM, Smith EE, Biessels GJ, et al. Neuroimaging standards for research into small vessel disease and its contribution to ageing and neurodegeneration. Lancet Neurol. 2013;12:822–838.

25. Vermeer SE, Den Heijer T, Koudstaal PJ, et al. Incidence and risk factors of silent brain infarcts in the population-based Rotterdam Scan Study. Stroke. 2003;34:392–396.

26. Longstreth WT, Jr., Arnold AM, Kuller LH, et al. Progression of magnetic resonance imaging-defined brain vascular disease predicts vascular events in elderly: the Cardiovascular Health Study. Stroke. 2011;42:2970–2972.

27. Vermeer SE, Prins ND, den Heijer T, et al. Silent brain infarcts and the risk of dementia and cognitive decline. N Engl J Med. 2003;348:1215–1222.

28. Hassell ME, Nijveldt R, Roos YB, et al. Silent cerebral infarcts associated with cardiac disease and procedures. Nat Rev Cardiol. 2013;10:696–706.

29. Schmidt R, Fazekas F, Offenbacher H, et al. Brain magnetic resonance imaging and neuropsychologic evaluation of patients with idiopathic dilated cardiomyopathy. Stroke. 1991;22:195–199.

30. Kozdag G, Ciftci E, Ural D, et al. Silent cerebral infarction in chronic heart failure: ischemic and nonischemic dilated cardiomyopathy. Vasc Health Risk Manag. 2008;4:463–469.

31. Siachos T, Vanbakel A, Feldman DS, et al. Silent strokes in patients with heart failure. J Card Fail. 2005;11:485–489.

32. Watson T, Shantsila E, Lip GY. Mechanisms of thrombogenesis in atrial fibrillation: Virchow’s triad revisited. Lancet. 2009;373:155–166.

33. Daniel WG, Nellessen U, Schröder E, et al. Left atrial spontaneous echo contrast in mitral valve disease: an indicator for an increased thromboembolic risk. J Am Coll Cardiol. 1988;11:1204–1211.

34. Merino A, Hauptman P, Badimon L, et al. Echocardiographic “smoke” is produced by an interaction of erythrocytes and plasma proteins modulated by shear forces. J Am Coll Cardiol. 1992;20:1661–1668.

35. Rodriguez Munoz D, Markl M, Moya Mur JL, et al. Intracardiac flow visualization: current status and future directions. Eur Heart J Cardiovasc Imaging. 2013;14:1029–1038.

36. Bermejo J, Martinez-Legazpi P, Alamo JCd. The clinical assessment of intracardiac flows. Ann Rev Fluid Mech. 2015;47:315–342.

37. Hong GR, Pedrizzetti G, Tonti G, et al. Characterization and quantification of vortex flow in the human left ventricle by contrast echocardiography using vector particle image velocimetry. JACC Cardiovasc Imaging. 2008;1:705–717.

38. Mangual JO, Domenichini F, Pedrizzetti G. Describing the highly three dimensional Right Ventricle flow. Ann Biomed Eng. 2012;40:1790–1801.

39. Kilner PJ, Yang G-Z, Wilkes AJ, et al. Asymmetric redirection of flow through the heart. Nature. 2000;404:759–761.

40. Martinez-Legazpi P, Bermejo J, Benito Y, et al. Contribution of the diastolic vortex ring to left ventricular filling. J Am Coll Cardiol. 2014;64:1711–1721.

41. Van Dantzig JM, Delemarre BJ, Bot H, et al. Doppler left ventricular flow pattern versus conventional predictors of left ventricular thrombus after acute myocardial infarction. J Am Coll Cardiol. 1995;25:1341–1346.

42. Asami R, Tanaka T, Kawabata KI, et al. Accuracy and limitations of vector flow mapping: left ventricular phantom validation using stereo particle image velocimetory. J Echocardiogr. 2017;15:57–66.

